# Discovery of Novel Trans-Ancestry and Ancestry-Specific Gene Loci for Total Testosterone in a Multi-Ancestral Analysis of Men in the Million Veteran Program

**DOI:** 10.1101/2022.02.16.21265846

**Authors:** Meghana S. Pagadala, Guneet K. Jasuja, Madhuri Palnati, Julie Lynch, Tori Anglin, Nelson Chang, Rishi Deka, Kyung Min Lee, Fatai Y. Agiri, Tyler M. Seibert, Brent S. Rose, Hannah Carter, Matthew S. Panizzon, Richard L. Hauger

## Abstract

Utilizing data from the Million Veteran Program (MVP), we investigated the genetic determinants underlying total testosterone levels via a multi-ancestral analysis of 124,593 individuals of European (n=88,385), African (n=25,235) and Hispanic (n=10,973) ancestry. We identified 46 trans-ancestry variants and 17 ancestry-specific variants, of which 14 trans-ancestry variants and 15 ancestry-specific variants are novel associations with testosterone. Results implicate genes regulating testosterone shared across ancestral groups, which include *SHBG, JMJD1C, FXR2, SENP3, TNFSF12-TNFSF13* while implicating genes such as *MSN, DMD, VSIG4, CHEK2, TKTL1* that may underlie ancestry-group differences in testosterone regulation. We also linked testosterone variants on the X chromosome with differential risk of chronic kidney disease and hereditary hemolytic anemias in African and Hispanic ancestry groups, respectively. Lastly, we constructed a polygenic score from our 46 trans-ancestry variants and associated it with testicular dysfunction, hyperlipidemia, gout and prostate cancer with stronger prostate cancer associations in Hispanic and African ancestry groups compared to the European ancestry group. These findings provide insight into ancestry-specific androgen regulation and identify novel variants for disease risk stratification in patients.

## Introduction

Testosterone is an anabolic steroid synthesized by the testis which acts as the primary sex hormone in men regulating sexual development during puberty and spermatogenesis and sexual function in adulthood^1–3^. Classical, well-established roles of testosterone include stimulation of erythropoiesis, maintenance of muscular strength, and maintenance of bone density mass;^4,5^ however, research has discovered that testicular androgens, testosterone and dihydrotestosterone have more extensive physiological actions with important roles in regulating cardiovascular, metabolic, hepatic, immune, and brain function^6–8^. These ubiquitous actions of testosterone are mediated by the androgen receptor (AR), encoded by the AR gene on the X chromosome, which is widely expressed in the testes, prostate, bone, skeletal muscle, heart, vascular smooth muscle, kidney, pulmonary epithelial cells, bone marrow hematopoietic and immune cells, adipose tissue, and the central nervous system^2,9^.

Beginning in midlife men experience a progressive reduction in testicular androgen steroidogenesis, which results in a roughly 1-3% decline in circulating levels of total testosterone per year^1,2,10,11^. Many men develop an age-related testosterone deficiency or hypogonadism, which contributes to sexual dysfunction, frailty, sarcopenia, coronary artery disease, metabolic syndrome and type 2 diabetes^1,2,10–12^. Testosterone levels in the hypogonadal range have been reported to increase the risk of prostate cancer and predict recurrence after prostatectomy and poor outcome in metastatic prostate cancer^13^. Testosterone deficiency has additionally been linked to depression, neurodegeneration, cognitive impairment, and Alzheimer’s disease risk during male aging^14–18^. To date, the genetic factors that contribute to individual differences in circulating testosterone levels in men during aging remain poorly understood. Furthermore, analysis of ancestry-specific genetic determinants governing testosterone regulation has not been studied.

Total testosterone levels are hecriptome-Wideritable (∼20%) and a number of genome-wide association studies (GWAS) of testosterone levels have been conducted, identifying strong associations with *SHBG, JMJD1C, FKBP4, REEP3*, and *FAM9B*^*19–25*^. However, most GWAS studies have been conducted primarily in European or Asian ancestry groups whereas differences in testosterone levels have been observed in men of other ancestries^26–28^. In this study, we conducted a GWAS of endogenous total testosterone levels in men enrolled in the Million Veteran Program (MVP), a large, multi-ethnic genetic biorepository from a hospital-based population. As a hospital-based population, MVP has higher disease prevalence of type 2 diabetes, obesity, hyperlipidemia, sleep apnea, gout, prostate cancer, and testicular dysfunction/hypofunction compared to other population studies and offers unique insights into testosterone regulation in a clinical setting. We identified 17 ancestry-specific testosterone gene variants and 46 trans-ancestry testosterone gene variants, of which 15 ancestry-specific variants and 14 trans-ancestry variants were novel. Colocalization and transcriptome-wide association study (TWAS) analysis revealed novel cell-type specific associations between *BRI3* and *BAIAP2L1*. Finally, we conducted phenome (PheWAS) and laboratory value association studies (LabWAS), which revealed shared associations of testosterone with type 2 diabetes, hyperlipidemia, gout and liver disease and ancestry-specific association with diabetes in the African ancestry group and hemolytic conditions in the Hispanic ancestry group. With this study, we gain insight into ancestry-specific regulation of testosterone, highlighting the importance of conducting future studies in diverse ancestry groups.

## Results

### Genome-wide analysis of Testosterone Levels

After phenotyping quality control, morning total testosterone levels were available for 145,576 male MVP participants (**Figure S1)**. MVP participants were categorized using Harmonized Ancestry and Race/Ethnicity (HARE), an algorithm which uses both self-reported and genetic ancestry to assign individuals to ancestry groups^29^ **(Table 1)**. Genome-wide association studies (GWAS) with testosterone levels were conducted separately in European (n=88,385), African (n=25,235) and Hispanic (n=10,973) ancestry groups. Incorporating all genetic information, we first assessed the heritability of testosterone levels from autosomal chromosomes across ancestry groups using LD score regression^30^. Total testosterone levels showed the highest heritability in the African ancestry group (h2=0.095) with significant GWAS hits (p<5e-08) accounting for 3.3% of heritability. Heritability of total testosterone levels in the European and Hispanic ancestry groups were 0.071 and 0.084 with significant hits (p<5e-08) accounting for 14.6% and 11.1% of heritability respectively **(Table S1)**. Variants passing the suggestive threshold (p<1e-05) explained 24.6%, 9.1%, 30.4% of heritability in European, African, and Hispanic ancestry groups, respectively.

**Table 1:** MVP Participant Characteristics for Testosterone GWAS. Table of MVP participants in European, African and Hispanic HARE groups used for testosterone GWAS.

|  | European | African | Hispanic |
| --- | --- | --- | --- |
| # male participants | 426,223 | 104,882 | 45,890 |
| # male participants with AM total testosterone levels | 88,385 | 25,235 | 10,973 |
| Mean total testosterone levels (Standard Deviation) ng/dl | 383 (189) | 408 (208) | 394 (189) |
| Mean age of testosterone lab (Standard Deviation) years | 60.45 (11.55) | 55.82 (9.93) | 54.72 (12.68) |

GWAS analyses identified 63 significant (p<5e-08) variants associated with testosterone across ancestry groups: 30 significant autosomal loci for the European ancestry group, 5 for the African ancestry group and 3 for the Hispanic ancestry group (**Figure 1)**. Several X chromosome variants remained significant after linkage-based clumping. To identify independent loci, we conducted a conditional analysis, where we used the most significant variants as covariates in the GWAS. Conditional analysis identified 9, 10 and 6 associations in European, African and Hispanic cohorts, respectively **(Figure 2A)**. Variants near *FAM9B, AR, RGAG1* were shared amongst all ancestry groups. X chromosome variant near *DMD* was identified specifically in the African ancestry group **(Figure 2B)**. *DMD* is responsible for dystrophin protein production and is mainly expressed in skeletal and cardiac muscle, although some expression in the brain has been observed^31–34^. A Hispanic ancestry group-specific variant near the ribosomal pseudogene *RPS29P28* was also identified **(Figure 2C)**.

**Figure 1:**
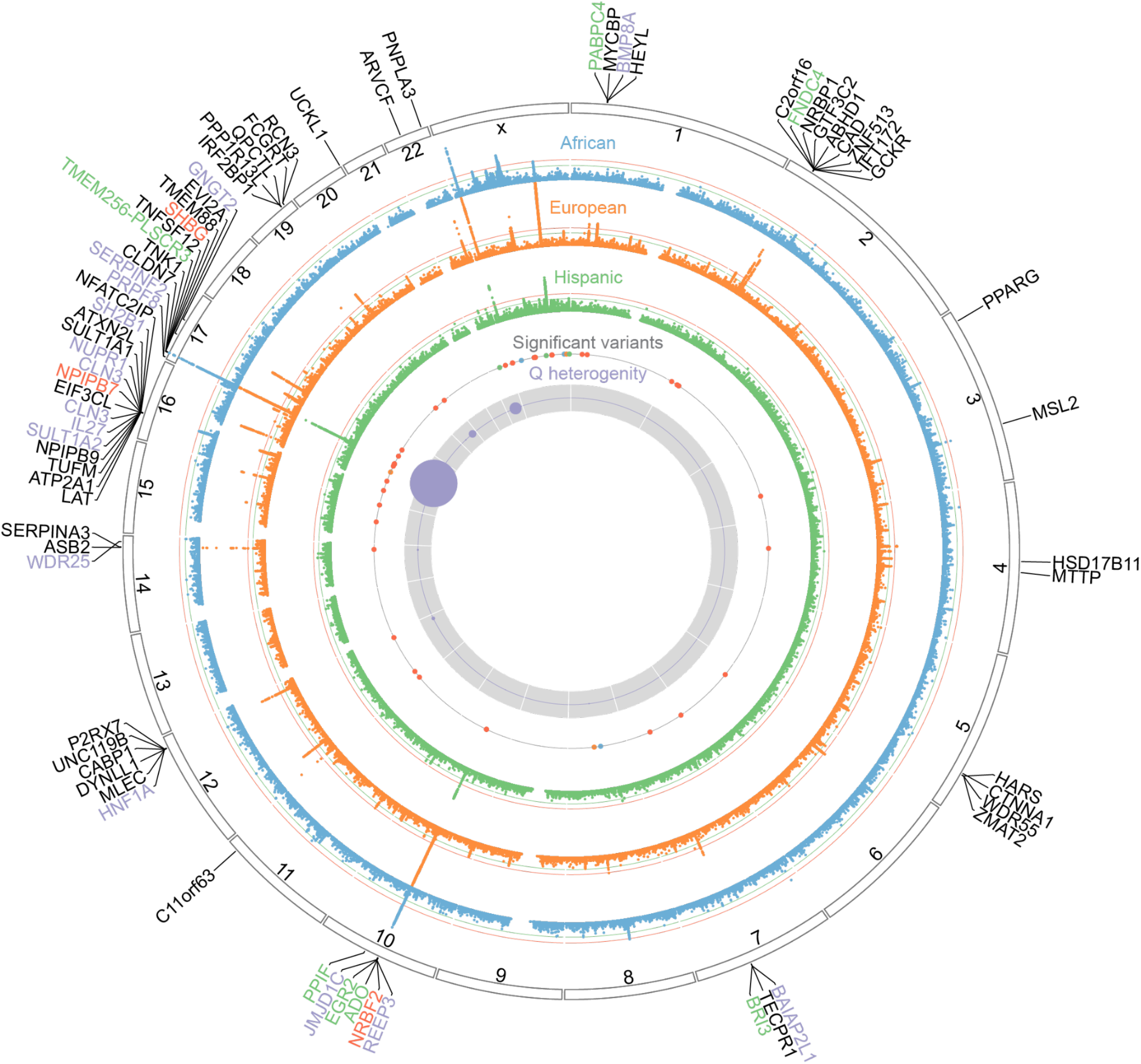
GWAS of Total Testosterone Levels in European, African and Hispanic HARE Groups. Circos plot of -log10 p values of Total Testosterone GWAS in European (orange), African (blue) and Hispanic (green) ancestry groups in the Million Veteran Project (MVP). The green line indicates the suggestive threshold (1e-06) and the red line indicates the genome-wide significance threshold (5e-08). The innermost purple band corresponds to measures of heterogeneity between ancestry group GWAS using Cochran’s Q test. Second innermost ring represents significant variants: red represents trans-ancestry variants, green represents Hispanic-specific variants, blue represents African-specific variants and orange represents European-specific variants. Selected genes near testosterone loci are indicated on the outside edge. A complete list of associations and nearby genes can be found in Supplemental Table 1.

**Figure 2:**
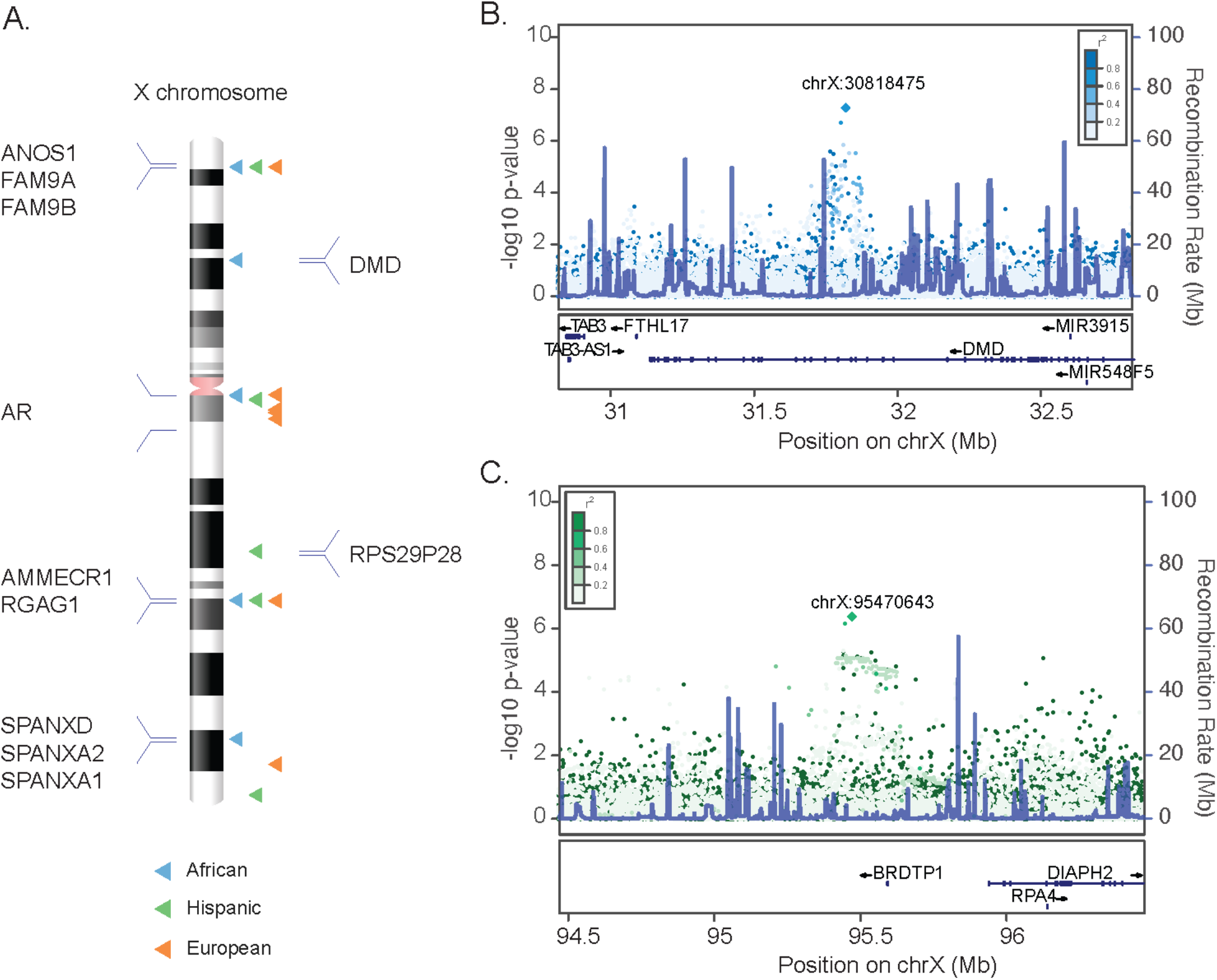
Sex Chromosome Analyses of Total Testosterone in European, African and Hispanic HARE Groups. **(A)** Karyotype plot of conditional X chromosome variants identified in European, African and Hispanic HARE groups. **(B)** Locuszoom plot of a conditional X chromosome variant in *DMD* uniquely identified in the African ancestry group. **(C)** Locuszoom plot of a conditional X chromosome variant near the *RPS29P28* pseudogene uniquely identified in the Hispanic ancestry group. A complete list of conditional associations can be found in Supplemental Table 2.

Loci on chromosomes 10 and 17 were shared across different ancestry groups. Most unique peaks were observed for the European ancestry group which had the largest sample size; however, unique peaks on chromosomes 3, 7, 8 and X were observed in the African ancestry group. We compared beta coefficients of SNP associations to determine if effects on testosterone were different amongst ancestry groups. Of the 63 variants, 10 variants had different effects on testosterone levels in the African ancestry group compared to the European ancestry group. Four of these variants were located on the X chromosome, with other variants located on chromosome 7, 8, 17, 19, and 22. Of the 63 variants, 8 variants had different effects on testosterone in the Hispanic ancestry group compared to the European ancestry group. Three of these variants were located on chromosome X, while the others were located on chromosomes 3, 7, 8, and 16. The chromosome 17 variant, rs980068134, was identified as a trans-ancestry variant and was associated with lower testosterone levels in African and Hispanic ancestry groups, but higher testosterone levels in the European ancestry group.

To validate the GWAS significant variants, we performed replication analyses for the European ancestry group variants in the UK Biobank. Of the 39 European ancestry group variants, 31 passed a significance threshold of 5e-08 in the UK Biobank (n=166,502) **(Figure S2A)**. Of the remaining 8 which did not pass GWAS significance threshold, 5 passed a significance threshold of 0.05. Comparable cohorts for the African and Hispanic ancestry group powered for replication were unavailable. However, we obtained total testosterone levels for individuals of African ancestry (n=1474) in the Multi-Ethnic Study of Atherosclerosis (MESA) study and found beta values of variants were concordant. **(Figure S2B)**.

### Genetic Correlation Across Ancestry Groups

To evaluate the extent to which variants modifying total testosterone levels are shared between ancestry groups, we estimated genetic correlation between groups by LD score (LDSC) regression^35^. While there was significant correlation between all groups, higher genetic correlation was observed between European and Hispanic ancestry groups (rg=0.60 [0.42-0.78]) compared to European and African ancestry groups (rg=0.19 [0.12-0.26]). Genetic correlation between African and Hispanic ancestry groups was 0.37 [0.16-0.58] **(Figure 3A)**.

**Figure 3:**
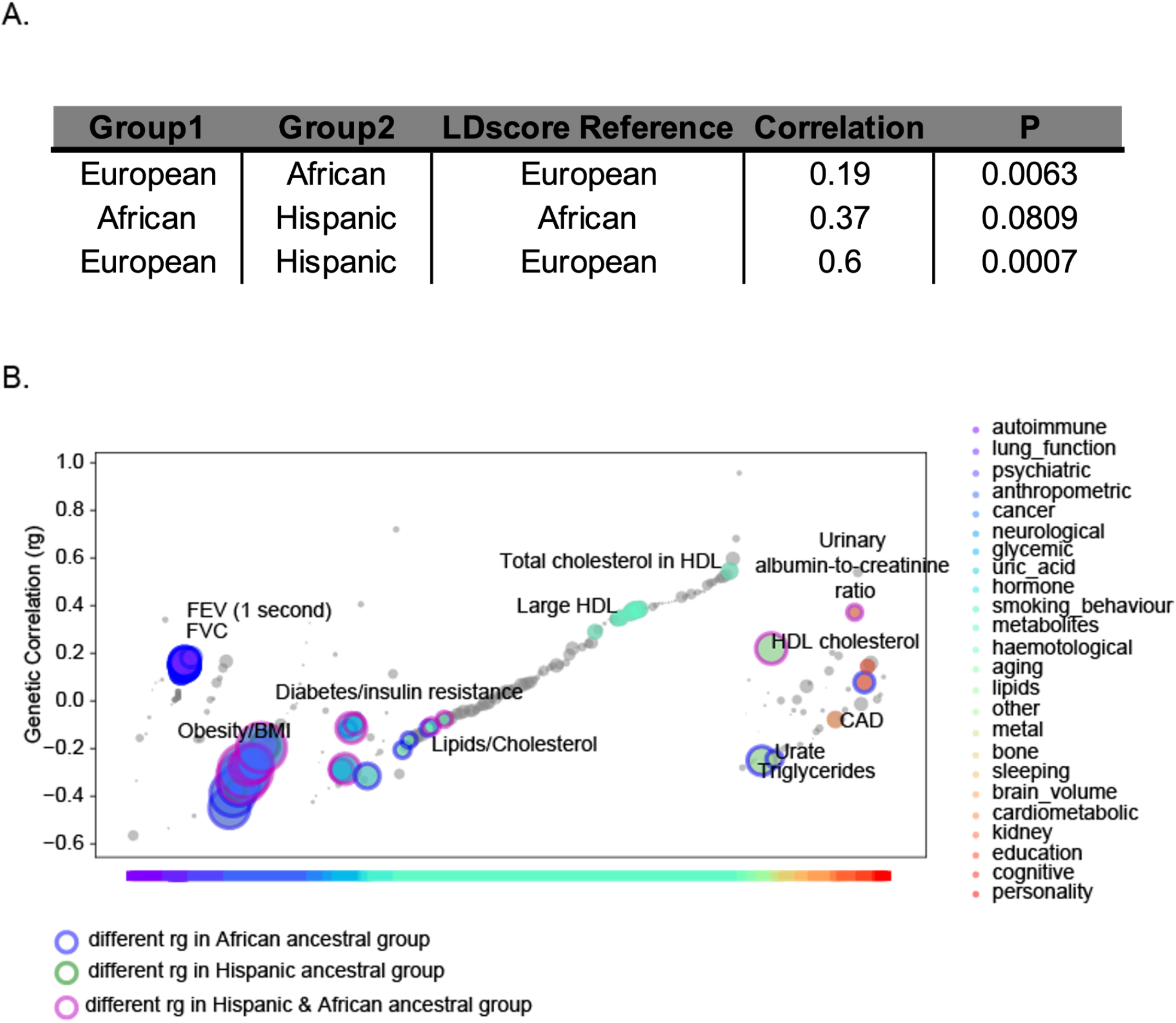
Genetic Correlation Across Ancestry Groups. **(A)** Genetic correlation between ancestry groups estimated by LSDC **(B)** Genetic correlation between testosterone and other traits from LDhub^36^ for European HARE groups. Genetic correlations different between ancestry groups are outlined in blue, if in African ancestry group only, green, if in Hispanic ancestry group only, and magenta is both in African and Hispanic ancestry groups. Selected categories are labels and full LDHub results are in Supplementary Tables 3-5.

We next conducted genetic correlation analysis between our total testosterone GWAS results and diseases/traits profiled in previous published GWASs using LDhub^36^, assessing differences in ancestry groups. Significant positive correlation with HDL and lung function, including forced vital capacity (FVC) and forced expiratory volume (FEV), were observed in European ancestry group (FDR < 0.1). Total testosterone levels were significantly negatively correlated with obesity, BMI, Type 2 Diabetes, triglycerides and VLDL (FDR < 0.1) in the European ancestry group. **(Figure 3B)**. No genetic correlations were significant in African and Hispanic ancestry groups; however, genetic correlations were highly concordant amongst ancestry groups **(Figure S3A)**. Positive correlations between testosterone and glucose were observed in the Hispanic ancestry group, but not European and African ancestry groups (**Figure S3B)**. These findings suggest potential differences in testosterone regulation of glucose between ancestry groups.

### Testosterone Variants Affect Gene Expression in Specific Cell Types

In order to identify candidate genes influencing testosterone levels we conducted a colocalization and a transcriptome-wide association study (TWAS) using GTEx gene expression information. The colocalization analysis estimates the probability of shared signal between *cis-*eQTL and GWAS analysis of testosterone^37^. Using a probability of colocalization > 0.8, we identified 28 genes modulating testosterone levels (**Figure 4)**. Tissue types with most identified genes included the thyroid (8) and aorta (6). Additionally, genes were identified in organs relevant to testosterone synthesis, such as the testis (3), prostate (2), liver (2) and adrenal gland (2). The colocalization analysis revealed high colocalization between testosterone variants and *cis* variants for *SHBG, TNFSF12, BRI3, LAT, NRBF2*, and *BAIAP2L1. SHBG* variants have been strongly associated with variation in testosterone action^38,39^. *BAIAP2L1* colocalization was specific to testes while *BRI3* colocalization was specific to the liver. Colocalization of *LAT* and *TNFSF12* was specific for the brain putamen/basal ganglia and brain spinal cord-cervical C1.

**Figure 4:**
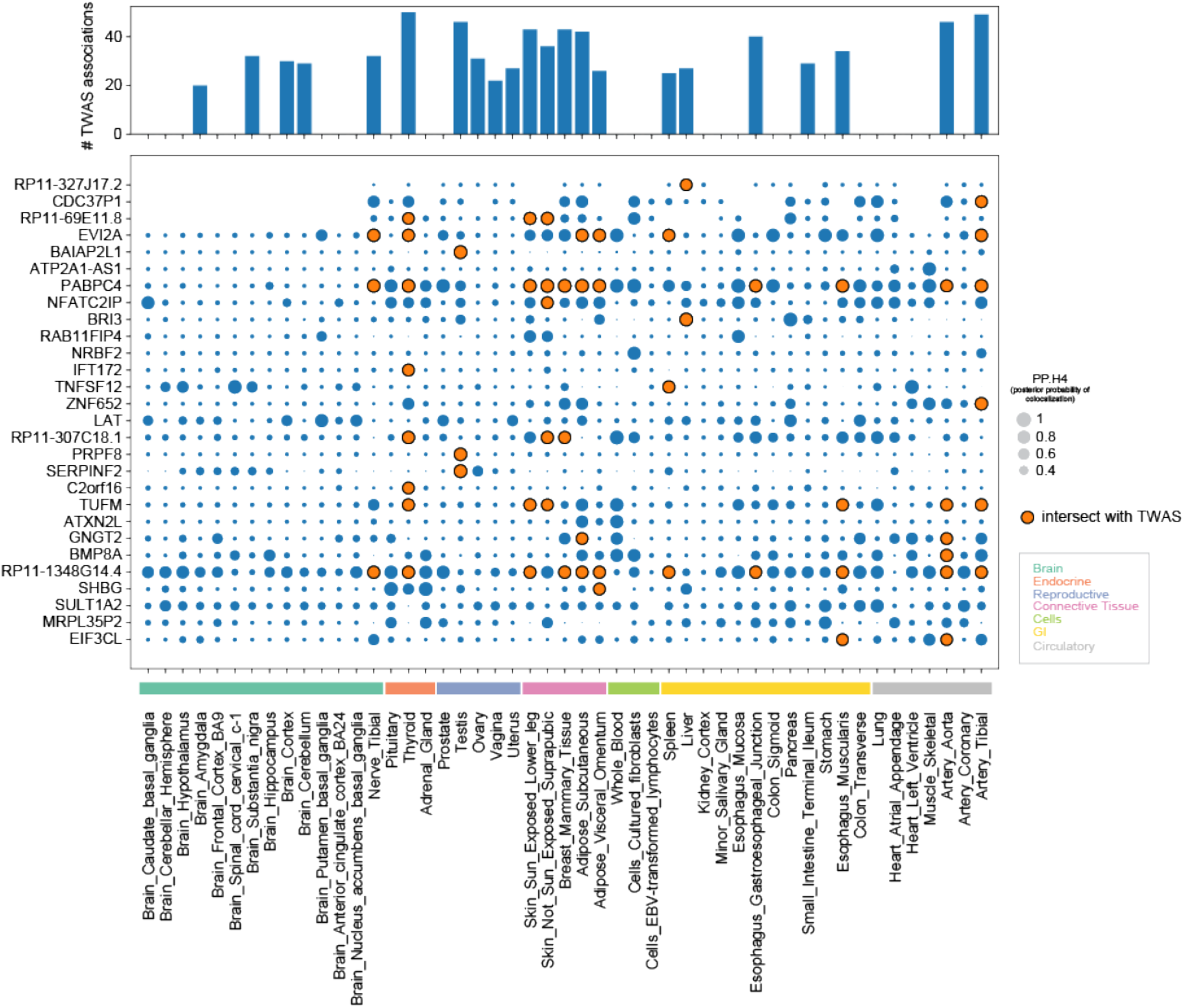
Testosterone Variants Affect Gene Expression in Specific Cell Types. Grid plot of genes with high colocalization (PP.H4 [posterior probability both traits are associated and share a single causal variant]>0.8) between Testosterone and GTEx *cis-*eQTL association by tissue type. Points in orange were also implicated by TWAS. Top barplot describes the number of TWAS associations for each cell type. Full colocalization and TWAS results are available in Supplemental Table 6 and 7, respectively.

The TWAS implicated 214 genes and 22 tissue types in total testosterone regulation. Tissue types for which most TWAS associations were identified included skin (sun-exposed), thyroid and tibial artery, consistent with tissue types identified through colocalization analysis. Forty-six associations with Testis and 27 associations with the Liver were observed. Of 28 genes identified by colocalization analysis, 23 were also identified by TWAS analysis. *BRI3* and *BAIAP2L1*, which are both located at chromosome 7, were identified specifically through liver and testis TWAS analysis, specifically. *BAIAP2L1* (Brain-Specific Angiogenesis Inhibitor 1-Associated Protein 2-Like Protein 1) is expressed in urothelial and glandular cells and has been implicated in previous GWAS studies of testosterone^40^. Gene expression studies identified *BAIAP2L1* to be upregulated with age and downregulated with riluzole treatment, a glutamate modulator shown to improve memory function in aged rats^41^. *BRI3*, or Brain protein I3, is a negative regulator of amyloid precursor protein (APP)^42^, providing a further link between the genetic determinants of testosterone regulation and Alzheimer’s disease pathophysiology. Although these genes have been implicated through tissues such as liver and testes, they might have broad effects on the human body and oncogenic mechanisms^43–45^.

To determine ancestry-associated differences in genetic regulation of testosterone, we conducted a Network Assisted Genomic Association (NAGA) on the GWAS results from each ancestry group and compared the top 50 implicated genes. Twenty-five genes were shared across all groups, including *SHBG, TNFSF12, SENP3, JMJD1C, RRAG1*, and *TP53*. Unique genes for the African ancestry group included *MSN, DMD, GAGE2B and GAGE2C*, while unique genes for the Hispanic ancestry group included *SERPINA7, TKTL1, TTC28*, and *PNPLA3*. Of the genes uniquely implicated, 9 and 2 were on chromosome X for African and Hispanic ancestry groups, respectively.

### PheWAS and LabWAS characterization of Testosterone Variants

Given the availability of extensive electronic health record data in the MVP, we conducted a phenome-wide association study (PheWAS) among individual level genotypes and ICD-10 code data to determine whether the 46 trans-ancestry testosterone variants were associated with disease risk. We identified 920, 45 and 56 significant associations (FDR < .01) in the European, African and Hispanic ancestry groups, respectively. Nine variants had PheWAS associations across all ancestry groups; 2 had associations uniquely in the African ancestry group. Across all ancestry groups, PheWAS associations for testicular dysfunction/hypofunction, hyperglyceridemia, gout, type 2 diabetes, chronic liver disease, hepatomegaly and hyperlipidemia were observed (**Figure 6A)**. In general, testosterone-increasing variants across ancestry groups were associated with lower odds of testicular hypofunction, dysfunction, morbid obesity, hyperglyceridemia, sleep apnea, diabetes mellitus, gout, hyperlipidemia and increased odds of hepatomegaly, liver disease and esophageal bleeding. Opposite to general trends, two variants on chromosome 2 and 17 (rs1260326, rs188272638) in *GCKR* and *POLR2A*, respectively, in the European ancestry group were associated with increased testosterone and increased obesity. Trans-ancestry variant rs190119169, an *AR* variant on chromosome X, was associated with increased testosterone across all ancestry groups (beta_eur=4.85, beta_afr=7.87, beta_his=6.11), but only significantly associated with lower odds of diabetes in the African ancestry group **(Figure 6B)**. Two testosterone-increasing variants, rs112635299 on chromosome 14 and rs764029425 on chromosome 7 near the *BAIAP2L1* gene, were significantly associated with lower odds of prostate cancer in the European ancestry group (rs112635299 OR=0.89 [0.86-0.91], rs764029425 OR=0.95 [0.94-0.96]).

**Figure 5:**
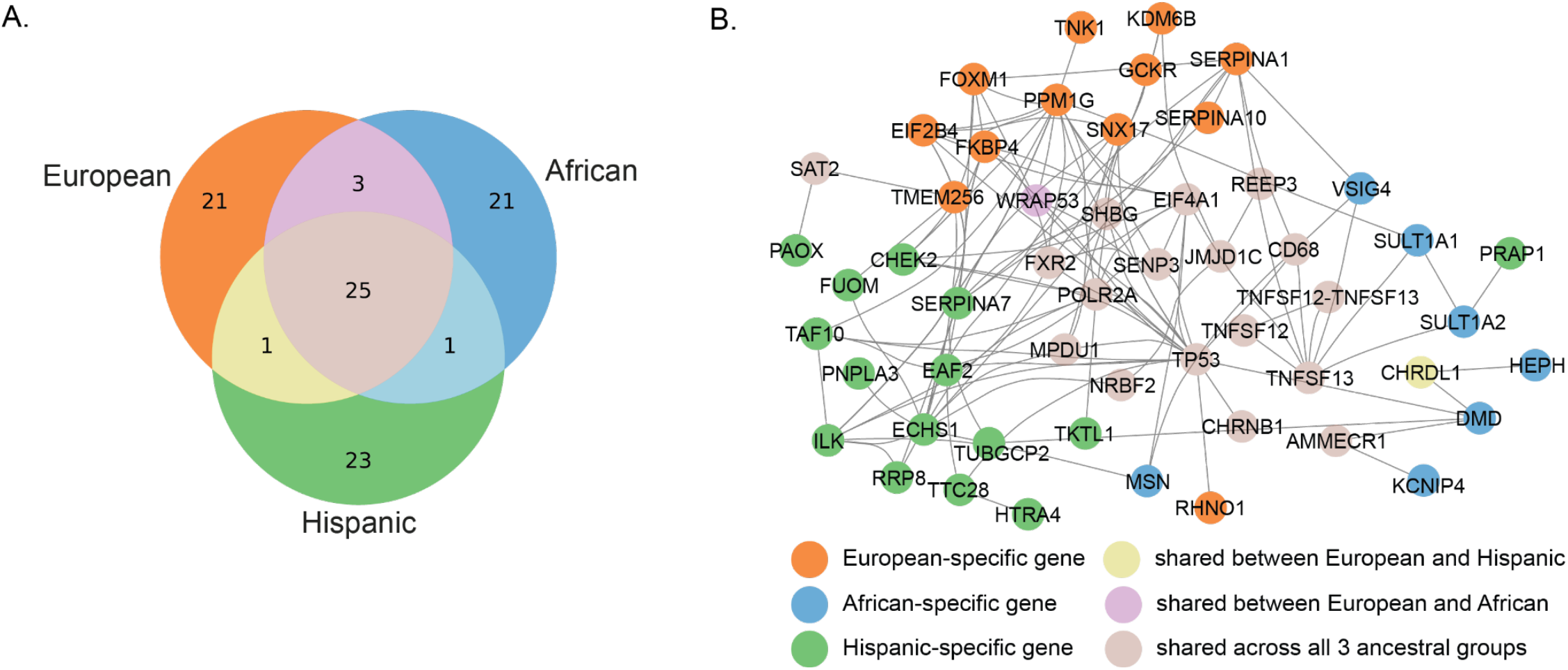
Shared Testosterone Genes Implicated by Network-Assisted Genomic Association (NAGA). **(A)** Venn diagram of top 50 genes implicated by NAGA analysis. **(B)** Fully connected subnetwork of genes implicated by NAGA. Nodes colored purple if shared across all 3 ancestry groups. European, African and Hispanic ancestry group-specific associations are colored orange, blue and green, respectively.

**Figure 6:**
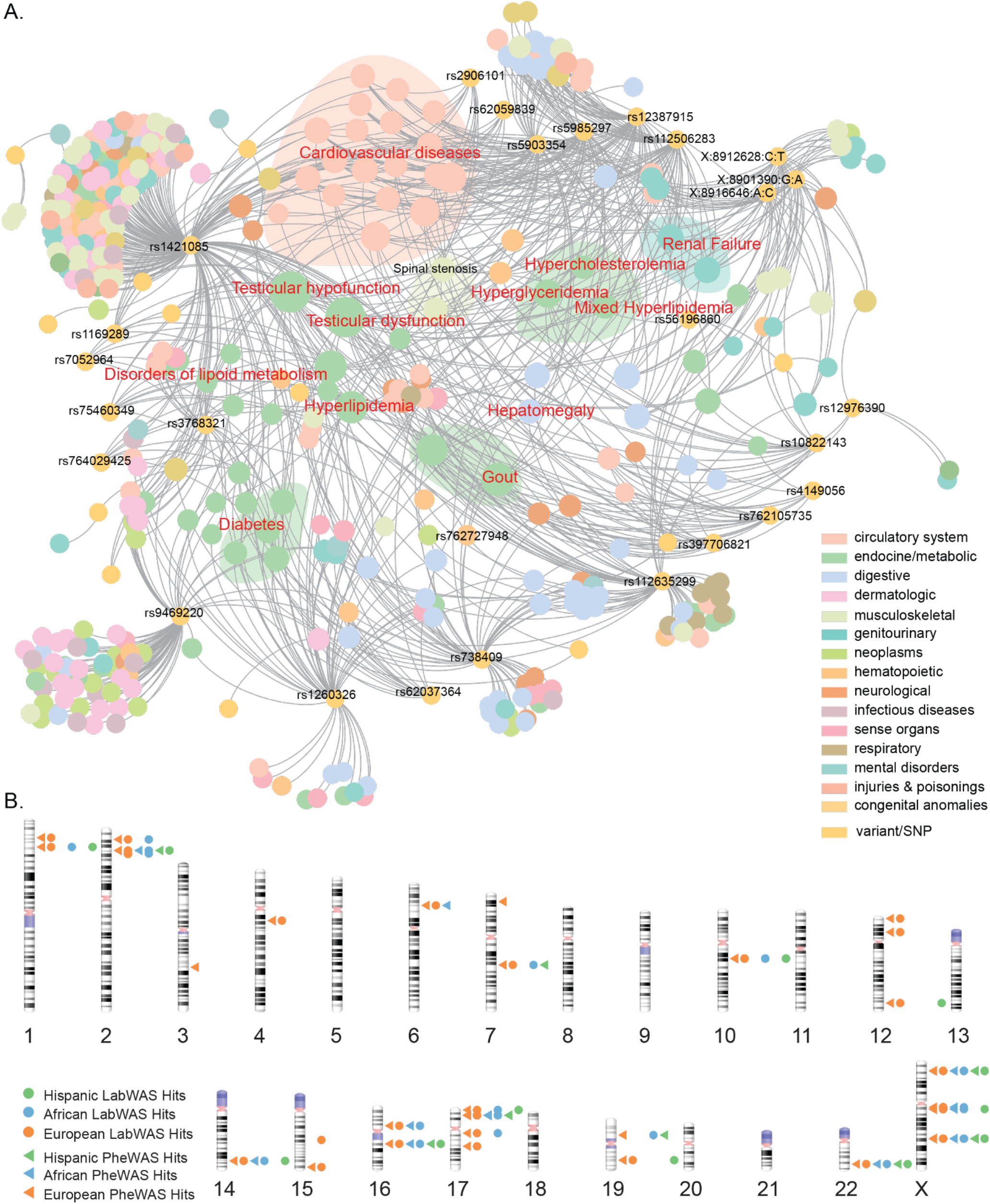
PheWAS associations highlight major diseases where testosterone genetic variants are implicated. **(A)** Gephi network plot of PheWAS associations in European ancestry group. **(B)** Karyotype plot of testosterone variants (red highlight) and any PheWAS association (triangles/circles). Full PheWAS and LabWAS results are found in Supplemental Tables 9-14.

Since testosterone can influence lab measurements^46^, we also ran LabWAS associations with 46 *trans-*ancestry variants and mean levels of lipid, metabolic and blood lab values. We identified 668 significant associations in the European, 150 in the African and 44 in the Hispanic ancestry groups (FDR<.01). Across all ancestry groups, significant associations with calcium, aspartate aminotransferase (AST), bicarbonate, A1c, hematocrit, estimated glomerular filtration rate (eGFR), alanine aminotransferase (ALT), HDL-C, iron, glucose, albumin, hemoglobin, LDL-C, international normalized ratio (INR), creatinine and chloride were observed. Of these shared associations, most associations with HDL-C, albumin and LDL-C were observed in the European ancestry group compared to glucose, hemoglobin and LDL-C in the African ancestry group. rs190119169, which was significantly associated with lower odds of diabetes in the African ancestry group, was also associated with significantly lower levels of mean glucose levels (beta=-5.14) in contrast to the European (beta=-0.60) and Hispanic (beta=-3.32) ancestry groups **(Figure 7A)**.

**Figure 7:**
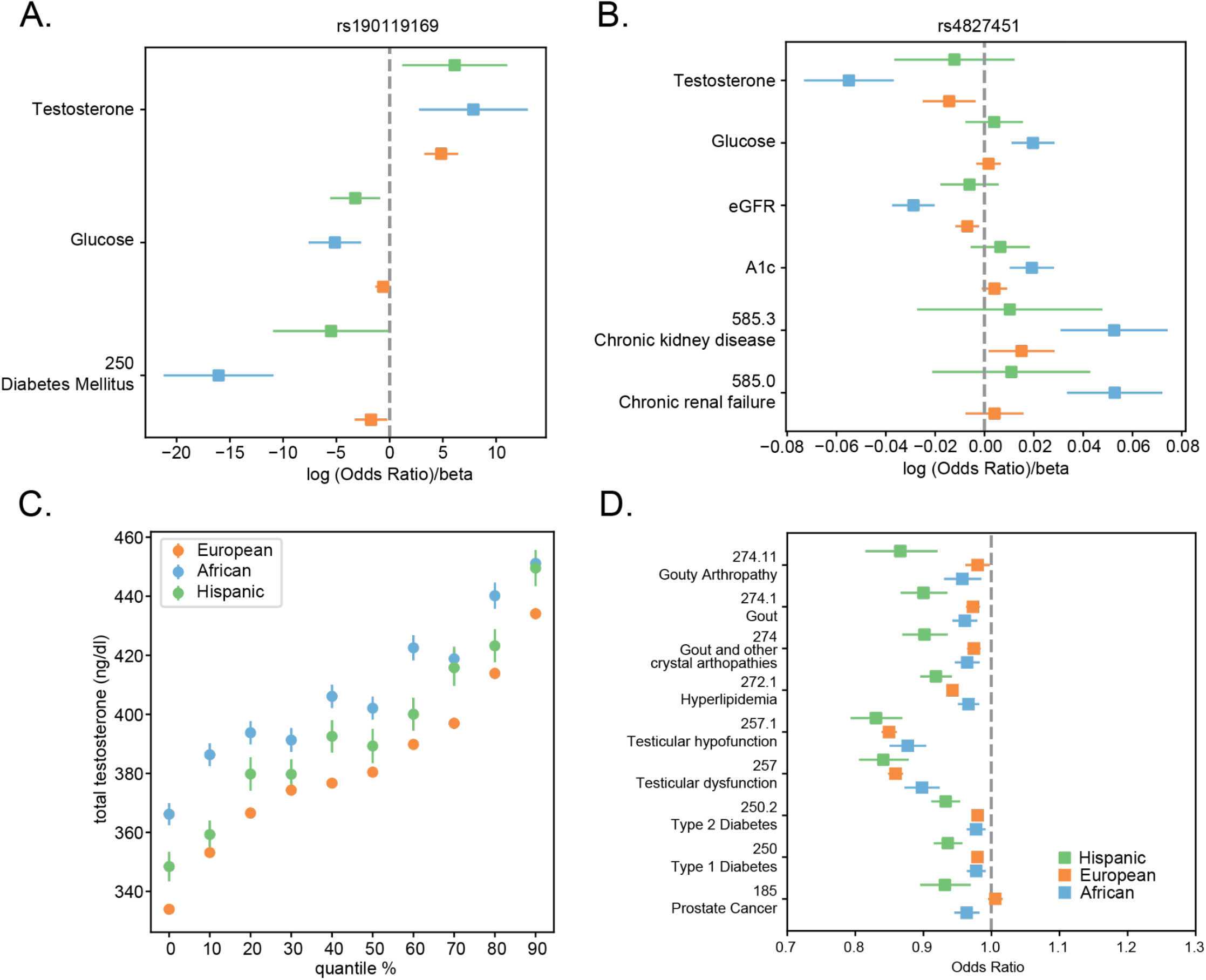
Ancestry-specific effects of Testosterone Variants and Polygenic Score (PGS). **(A)** Plot of chromosome X variant rs190119169 ancestry-specific effects on testosterone levels, average glucose lab levels and diabetes risk. **(B)** Plot of chromosome X variant rs4827451 ancestry-specific effects on testosterone levels, average glucose levels, average eGFR, A1c levels and chronic kidney disease and renal failure risk. **(C)** Quantile plot of testosterone PGS and testosterone levels (ng/dl) in the MVP discovery cohort. **(D)** Odds ratio plot of the testosterone PGS association with testicular dysfunction, testicular hypofunction, hyperlipidemia, gout, type 1 and 2 diabetes, and prostate cancer.

We next evaluated PheWAS and LabWAS associations of 17 ancestry-specific variants. One of the 8 African-ancestry specific variants and 3 of the 5 Hispanic ancestry-specific variants had PheWAS association in respective ancestry groups. African-specific ancestry variant rs4827451 on chromosome X was uniquely associated with diabetes, diabetic retinopathy and renal failure in the African ancestry group. This variant was also uniquely associated with A1c, glucose and eGFR in the African ancestry group **(Figure 7B)**. One of the genes in close proximity to rs4827451 is *MSN*, which could have potential mechanisms in renal fibrosis. rs73629199, a *TKTL1* variant on chromosome X, was uniquely associated with hereditary hemolytic anemias in the Hispanic ancestry group. Furthermore, significant associations were observed with hematocrit, ferritin, hemoglobin in the Hispanic ancestry group along with the African ancestry group.

### Testosterone Polygenic Score (PGS) is predictive of Testicular Dysfunction and Hyperlipidemia Across All Ancestry Groups

Given the numerous PheWAS and LabWAS associations with testosterone variants, we wanted to assess the predictive value of testosterone variants for different clinical phenotypes. We constructed a testosterone polygenic score from 46 trans-ancestry variants, which we validated in the MVP cohort **(Figure 7C)** and the UK Biobank (**Figure S4)**. Distribution of testosterone polygenic scores amongst ancestry groups were significantly different (t-test p < 2e-16). The African ancestry group had the highest average testosterone PGS scores (mean=7.76, std=0.26) followed by the European (mean=5.88,std=0.34) and Hispanic ancestry groups (mean=4.69, std=0.40) **(Figure S5)**.

To assess the predictive value of the testosterone PGS for clinical phenotypes, we conducted association analyses between the testosterone PGS and the clinical phenotypes with PheWAS associations across ancestry group, namely testicular dysfunction, testicular hypofunction, hyperlipidemia, gout, Type 1 and 2 diabetes, and prostate cancer. The testosterone PGS was associated with all clinical phenotypes with the strongest effects observed for testicular dysfunction (OR_eur_ = 0.86 [0.85-0.87], OR_afr_ = 0.90 [0.87-0.92], OR_his_ = 0.84 [0.81-0.88]) and testicular hypofunction (OR_eur_=0.85 [0.84-0.86], OR_afr_=0.88 [0.85-0.90], OR_his_ = 0.84 [0.79-0.87]). Higher testosterone PGS was associated with lower odds of gout, hyperlipidemia, Type 1 and 2 diabetes, and prostate cancer **(Figure 7D)**. Higher testosterone PGS was associated with significantly lower odds of gout among Hispanics as compared to the European and African ancestry groups. Furthermore, testosterone PGS was uniquely associated with lower odds of prostate cancer in the Hispanic and African ancestry groups, but not European ancestry group. These results suggest a differential role for testosterone regulation in disease risk stratification across ancestry groups.

## Discussion

Utilizing data from the MVP cohort, we present the largest multi-ancestral genome-wide analysis of total testosterone levels to our knowledge. We identified 46 trans-ancestry variants of which 14 were novel. We identified 17 ancestry-specific variants in European (8), African (5), and Hispanic (4) ancestries of which 15 were novel. Of the 63 testosterone variants, 32 trans-ancestry and 2 ancestry-specific variants were in high LD (R2 > 0.2) with variants identified in UK Biobank analysis of testosterone levels^19^. The genes implicated by shared variants include *SAT2, SHBG, FXR2, MPDU1, SOX15, TNFSF12-TNFSF13, SENP3, FGF11, CHRNB1, ZBTB4, POLR2A, TP53, JMJD1C, NRBF2, TDGF1P3, RGAG1*, and *AMMECR1. MSN, DMD, VSIG4*, and *HEPH* were genes implicated by African ancestry group analysis specifically. *CHEK2, TTC28*, and *TKTL1* were genes implicated by Hispanic ancestry group analysis.

It is well-established that testosterone is immunosuppressive dampening many aspects of humoral-and cell-mediated immunity. Interestingly, both *VSIG4* and *MSN*, implicated uniquely by African ancestry group analysis, are immunomodulators. *VSIG4* is a macrophage-expressed complement receptor of the immunoglobulin superfamily with T-cell suppressive and differentiation effects^47–49^ while *MSN* is an adaptor molecule essential for cell shape, motility and signaling regulation^50–52^. *MSN* improves prognosis in lung adenocarcinoma patients by enhancing immune lymphocyte infiltration^53^. Studies have linked testosterone with *MSN* expression, demonstrating the interaction promotes actin cytoskeletal remodeling and enhanced migration in endothelial cells^54^. The immunomodulatory effects of testosterone have downstream effects for atherosclerosis, COVID-19 infection, and other diseases^55,56^. Identification of immunomodulatory genes specifically in the African ancestry group suggests an ancestry-specific genetic link between testosterone and the immune system that is important to explore.

Our analysis found novel European ancestry-specific variants for the Ephrin B1 (EFNB1) gene at Xq13 and the WW Domain Containing Oxidoreductase (WWOX) gene at 16q23.1-2 that were associated with testosterone levels. The *EFNB1* gene encodes a ligand for Ephrin receptor tyrosine kinases, which has critical roles in migration and adhesion of brain neurons and cells in the lung, adipose tissue, kidney, and cardiovascular system during development. Recently, EFNB1 in hippocampal astrocytes has been found to regulate excitatory and inhibitory neurocircuits and exert a neuroprotective role in the adult brain^57,58^. The WWOX protein contains a short–chain dehydrogenase reductase (SDR) domain at its C-terminus that is involved in androgen steroidogenesis in Leydig cells of the testis; knockout of the WWOX results in a severe testosterone deficiency, impaired hematopoiesis and cholesterol metabolism, and renal failure^59^. Furthermore, a recent genetic meta-analysis discovered that the WWOX locus has a genome-wide association with Alzheimer’s disease^60^. Low testosterone levels in men is a risk factor for dementia^61^. Furthermore, testosterone replacement therapy (TRT) can modestly improve cognition^62^, suggesting testosterone has a potential protective effect for dementia. Novel testosterone loci in genetic regions relevant to dementia may be due to the higher representation of disease in the MVP that are not available in other biobanks and highlight genetic links between testosterone and memory function.

The *SHBG* locus on chromosome 17 was significantly associated with testosterone levels across ancestry groups, however, differences in PheWAS associations at this locus were observed in the three ancestries. rs918121801 was only associated with testicular dysfunction and hypofunction in the African ancestry group while rs62059839, in close proximity to rs918121801, was associated with testicular dysfunction and hypofunction in the European ancestry group. High heterogeneity at the *SHBG* locus was calculated amongst ancestry groups and *SHBG* levels are known to be highly heritable^63–65^. Thus, differences in PheWAS associations within the *SHBG* locus may reflect complex linkage disequilibrium patterns between ancestry groups^66^ or a shared causal variant with different ancestry group specific markers. Causality analysis and fine mapping of the *SHBG* locus may help to better understand ancestry-specific differences in testosterone regulation.

We also linked ancestry-specific variants with differential disease risk in African and Hispanic ancestry groups. Specifically, chromosome X variant rs4827451 was associated with lower testosterone levels and eGFR and higher odds of chronic kidney disease and renal failure. Hypogonadism is frequently observed in renal failure^67^, increasing risk of anemia and cardiovascular disease. Furthermore, sex-specific and ancestry-specific disparities in CKD are important, with kidney function deteriorating faster in men compared to women and higher rates of renal failure occurring in African Americans compared to Caucasians. Pathophysiological mechanisms underlying renal failure are complex^68^, but maintaining androgen regulation of renal function may have a therapeutic effect on chronic kidney disease improving medical management^69^. Our identified variant suggests a potential genetic mechanism linking hypogonadism and chronic kidney disease in individuals of African ancestry group specifically. Furthermore, the variant is located in close proximity to *MSN*, which is involved in renal fibrosis through E-cadherin interaction^70^. These findings demonstrate a potential ancestry-specific genetic mechanism in the kidney that requires further investigation.

A unique PheWAS association between a *TKTL1* variant and hereditary hemolytic anemias was identified in the Hispanic ancestry group. Lower testosterone levels are associated with higher rates of anemia in men, while testosterone replacement therapy stimulates iron-dependent erythropoiesis and has beneficial effects on anemia. Given the high heritability of red blood cell (RBC) traits^71^, the identified *TKTL1* variant may give insight into androgen-related genetic regulation of anemia in non-European individuals. Furthermore, rs73629199 was associated with RBC-related lab values such as ferritin, hematocrit, and hemoglobin that links testosterone with RBC regulation.

Of the 17 ancestry-specific variants, 8 were identified in the African, 5 were identified in Hispanic, and 4 were identified in European ancestry group analyses. Most African ancestry-specific variants were found on chromosome X, however, we discovered *LIN01414* was a unique variant on chromosome 8. Based on LD score regression analysis, heritability of testosterone levels were highest in the African ancestry group; however, significant variants only explain a minority of heritability (3.3%). Even using a suggestive threshold, heritability explained was 9.1% compared to 24.6% and 30.4% in European and Hispanic ancestry groups, respectively. Differences in total testosterone levels have been observed in African individuals and several of the diseases implicated by PheWAS analysis of testosterone levels, such as diabetes, cardiovascular disease, and chronic renal disease, occur at higher rates in individuals of African ancestry^72–81^. Understanding androgen-related regulation of diseases in individuals of African ancestry is critical and will require larger sample sizes.

We constructed a polygenic score (PGS) for testosterone using 46 trans-ancestry variants and observed differential associations with gout and diabetes in the Hispanic ancestry group compared to European and African ancestry groups. Furthermore, PGS was associated with lower odds of prostate cancer in the Hispanic and African ancestry groups compared to the European ancestry group. In hyperlipidemia and testicular dysfunction, PGS associations were more concordant across ancestry groups. These findings highlight shared effects of testosterone on disease risk, but also reveal differences that give insight into ancestry group-specific genetic architecture of testosterone.

The Million Veteran Program provides a breadth of clinical and genomic data that has allowed us to conduct the most ancestrally diverse genetic analysis of total testosterone levels to date. We note limitations with exploring ancestry group-specific mechanisms as the tissue-specific gene expression data that are available come predominantly from individuals with European ancestry. Thus, our TWAS and colocalization analysis rely on European gene expression data; however, gene expression patterns may be influenced by ancestry group differences. Furthermore, our PheWAS analysis focused on ICD10 codes for clinical phenotyping. These clinical outcomes are important for assessing disease risk; however, testosterone can also impact disease severity, treatment response and disease progression. Future directions should include continuing clinical phenotyping efforts in the MVP and assessing interactions with genomic determinants of testosterone and its impact of health and disease.

In summary, we conducted a large-scale multi-ancestry GWAS of morning total testosterone levels in men from the Million Veteran Program and discovered multiple novel trans-ancestry and ancestry-specific variants influencing testosterone levels. Using additional clinical and gene expression data, we linked ancestry-group germline variant differences with differential disease risk and constructed a polygenic risk score predictive of a variety of clinical phenotypes. These results provide unique insights into not only testosterone regulation but also ancestry group differences in genetic regulation of testosterone.

## Methods

### Genome-wide Association Analysis

For Million Veteran Program (MVP) genome-wide association studies (GWAS), total testosterone levels (ng/dl) between 7AM and 12PM for 88,385 individuals of European ancestry, 25,235 individuals of African ancestry and 10,973 individuals of Hispanic ancestry as determined by HARE groups were inverse rank normalized. PLINK glm method was applied to conduct association analyses with testosterone levels excluding variants of minor allele frequency threshold less than 0.1% and correcting for covariates: principal components (1-10), age. Patients with X chromosome aneuploidy (XXY, XYY) were excluded from analysis. Patients on androgen deprivation therapy (ADT), and testosterone replacement therapy (TRT) were excluded from analysis to focus on analysis of endogenous testosterone levels. In patients with multiple testosterone levels readings, first testosterone reading was used.

For European ancestral group testosterone loci validation, field ID 30850 (Testosterone nmol/L) was extracted and inverse-rank normalized from UK Biobank. PLINK^82^ glm method was used to calculate beta and p-values for variants identified in MVP discovery analysis. UK Biobank validation cohort consisted of 166,502 European, unrelated men. Top 10 principal components (PC1-PC10) and age were used as covariates.

For African ancestral group testosterone loci validation, total testosterone values were extracted and inverse-rank normalized for self-identifying African individuals in the Multi-Ethnic Study of Atherosclerosis (MESA). Individuals with KING relatedness > 0.177 were removed. PLINK glm method was used to conduct validation analysis of testosterone loci in 1317 men. Age was used as a covariate.

### Conditional X chromosome analysis

PLINKv2.0 glm method was used to run stepwise conditional analysis for identification of independent X chromosome associations^83^. First, association with testosterone was conducted and SNP with lowest p-value was identified. Next association with testosterone was run conditioning on most significant variants. Analysis was conducted until no SNPs with p-value < 1×10^−6^ remained. To reduce the chance of multicollinearity, variants which were within 50kb of the most recent significant variant were excluded from the selection process.

### LDSC

LD scores for European, African and Hispanic ancestral groups were calculated using LDSC^30^ from 1000 Genomes EUR, AFR, and AMR groups, respectively. For partitioned heritability analysis, categories for analysis included base and significant category for variants which based p-value threshold (p < 5e-08) for each ancestral group^84^. Genetic correlation analysis was conducted with summary statistics for each ancestral group using LD scores from both groups in analysis^35^. Lastly, for LD hub analysis, summary statistics were uploaded to LD Hub server (http://ldsc.broadinstitute.org/). Genetic correlations in UK Biobanks and “reproductive” category were excluded. FDR was calculated from LD hub provided p-values.

### Colocalization Analysis

Significant variants (p < 5e-08) for each ancestral group were compared with significant GTEx eQTL results (accession date: 10/02/2019). All genes for which significant eQTL association was shared were included in colocalization analysis. Full summary statistics for GTEx *cis*-eQTL analysis were used to make input files including samples sizes and p-values for both GTEx and MVP testosterone summary statistics from European ancestral group. Minor allele frequency from GTEx consortium was included for final coloc analysis. coloc.abf function in coloc R package^37^ was used to conduct analysis. Genes with PP.H4 (posterior probability of one common causal variant between studies) of 0.8 were analyzed.

### TWAS

TWAS R package was downloaded from TWAS hub using 1000 genomes PLINK files to infer linkage disequilibrium patterns and 22 GTEx expression models^85^. TWAS analysis was conducted with all expression models using European linkage-disequilibrium reference from 1000 Genomes.

### PheWAS & LabWAS

1817 International Classification of Diagnoses-10-Clinical Modification (ICD-10 CM) diagnosis codes from electronic health care records were available for MVP participants from as early as 1998. PheWAS R package was used to conduct PheWAS associations in male MVP European (n=421,212), African (n=104,380), Hispanic (n=45,553) ancestral groups. Cases were individuals with investigated phecode, whereas controls did not have incidence of phecode. Covariates included age at testosterone lab and first 10 genetic principal components.

Mean lab values for 68 measurements were available. As was done for the PheWAS, associations with mean lab values were conducted with PLINK using top 10 principal components and age as covariates.

False discovery rates (FDR) for PheWAS and LabWAS associations were calculated using Benjamini-Hochberg correction^86^ with python statsmodel package^87^. Associations with FDR less than 1% were considered significant.

### Testosterone Polygenic Score (PGS)

Meta-analysis of European, African and Hispanic testosterone GWAS were conducted with METAL. Variants passing significance threshold (p<5e-08) in meta-analysis were used to construct polygenic scores. 46 trans-ancestry variants were extracted from imputed genotype PGEN files in Million Veteran Program (MVP). Alleles were oriented to increasing testosterone levels. Genotype dosages were weighted by beta values for each respective ancestry group and summed. Logistic regression analyses with phecodes extracted from PheWAS dataframe were conducted with age of enrollment and top 10 principal components as covariates. For UK Biobank validation, only 37 of the 46 variants were found and used to construct testosterone PGS. Dosages were weighted by beta values from MVP European ancestry group analysis.

## Data Availability

Requests regarding data access may be directed to.

## Acknowledgements

This research used data from the Million Veteran Program, Office of Research and Development, Veterans Health Administration. This research was supported by the Million Veteran Program MVP022 award # I01 CX001727 (PI: Richard L. Hauger MD). This publication does not represent the views of the Department of Veterans Affairs or the United States Government. Dr. Hauger was additionally funded by the VISN-22 VA Center of Excellence for Stress and Mental Health (CESAMH) and National Institute of Aging RO1 grant AG050595 (*The VETSA Longitudinal Twin Study of Cognition and Aging VETSA 4)*.

For LDhub analysis, we gratefully acknowledge all the studies and databases that made GWAS summary data available: ADIPOGen (Adiponectin genetics consortium), C4D (Coronary Artery Disease Genetics Consortium), CARDIoGRAM (Coronary ARtery DIsease Genome wide Replication and Meta-analysis), CKDGen (Chronic Kidney Disease Genetics consortium), dbGAP (database of Genotypes and Phenotypes), DIAGRAM (DIAbetes Genetics Replication And Meta-analysis), ENIGMA (Enhancing Neuro Imaging Genetics through Meta Analysis), EAGLE (EArly Genetics & Lifecourse Epidemiology Eczema Consortium, excluding 23andMe), EGG (Early Growth Genetics Consortium), GABRIEL (A Multidisciplinary Study to Identify the Genetic and Environmental Causes of Asthma in the European Community), GCAN (Genetic Consortium for Anorexia Nervosa), GEFOS (GEnetic Factors for OSteoporosis Consortium), GIANT (Genetic Investigation of ANthropometric Traits), GIS (Genetics of Iron Status consortium), GLGC (Global Lipids Genetics Consortium), GPC (Genetics of Personality Consortium), GUGC (Global Urate and Gout consortium), HaemGen (haemotological and platelet traits genetics consortium), HRgene (Heart Rate consortium), IIBDGC (International Inflammatory Bowel Disease Genetics Consortium), ILCCO (International Lung Cancer Consortium), IMSGC (International Multiple Sclerosis Genetic Consortium), MAGIC (Meta-Analyses of Glucose and Insulin-related traits Consortium), MESA (Multi-Ethnic Study of Atherosclerosis), PGC (Psychiatric Genomics Consortium), Project MinE consortium, ReproGen (Reproductive Genetics Consortium), SSGAC (Social Science Genetics Association Consortium) and TAG (Tobacco and Genetics Consortium), TRICL (Transdisciplinary Research in Cancer of the Lung consortium), UK Biobank. We gratefully acknowledge the contributions of Alkes Price (the systemic lupus erythematosus GWAS and primary biliary cirrhosis GWAS) and Johannes Kettunen (lipids metabolites GWAS).

## Author Contributions

M.P. and R.H. conceived the work and designed the experiments; M.P., N.C., R.D., K.L., T.A., and J.L. assisted in clinical information processing; M.S.P. performed GWAS, PheWAS, and LabWAS analysis; M.S.P. wrote the paper with assistance from G.K., T.S., B.R., J.L., H.C., M.P., R.H..; G.K., T.S., B.R., J.L., H.C., M.P., and R.H. advised on genetic analysis

## Data Availability

The full summary-level association data from this report are available upon request. Source data are provided with this paper.

## Code Availability

Code to reproduce manuscript figures is available at: https://github.com/meghanasp21/mvp-testosterone-gwas

## Supplementary Tables

**Supplemental Table 1** 63 Trans-Ancestry and Ancestry-Specific Associations with Total Testosterone in Million Veteran Program (MVP)

**Supplemental Table 2** Conditional Chromosome X Associations with Total Testosterone in Million Veteran Program (MVP)

**Supplemental Table 3** Colocalization Results for Total Testosterone and GTEx eQTL information for 49 Tissues

**Supplemental Table 4** Transcriptome-Wide Association Study (TWAS) Results for Total Testosterone and GTEx Weight Matrices for 48 Tissues

**Supplemental Table 5** Disease Prevalence Differences for 16 ICD-10 Phecodes Across UK Biobank and Million Veteran Program (MVP)

**Supplemental Table 6** PheWAS Association Results for European HARE group in Million Veteran Program (MVP)

**Supplemental Table 7** PheWAS Association Results for African HARE group in Million Veteran Program (MVP)

**Supplemental Table 8** PheWAS Association Results for Hispanic HARE group in Million Veteran Program (MVP)

**Supplemental Table 9** LabWAS Association Results for European HARE group in Million Veteran Program (MVP)

**Supplemental Table 10** LabWAS Association Results for African HARE group in Million Veteran Program (MVP)

**Supplemental Table 11** LabWAS Association Results for Hispanic HARE group in Million Veteran Program (MVP)

**Supplemental Table 12** LDHub Results with for European HARE group in Million Veteran Program (MVP)

**Supplemental Table 13** LDHub Results with for African HARE group in Million Veteran Program (MVP)

**Supplemental Table 14** LDHub Results with for Hispanic HARE group in Million Veteran Program (MVP)

## References

1. Bhasin, S. et al. Testosterone Therapy in Men With Hypogonadism: An Endocrine Society Clinical Practice Guideline. J. Clin. Endocrinol. Metab. 103, 1715–1744 (2018).

2. Matsumoto, A. M. & Bremner, W. J. Testicular Disorders. Williams Textbook of Endocrinology 688–777 (2011) doi:10.1016/b978-1-4377-0324-5.00019-5.

3. Wilson, J. D., Griffin, J. E., WGeorge, F. & Leshin, M. The Endocrine Control of Male Phenotypic Development. Australian Journal of Biological Sciences vol. 36 101 (1983).

4. Shahani, S., Braga-Basaria, M., Maggio, M. & Basaria, S. Androgens and erythropoiesis: past and present. J. Endocrinol. Invest. 32, 704–716 (2009).

5. Herbst, K. L. & Bhasin, S. Testosterone action on skeletal muscle. Current Opinion in Clinical Nutrition and Metabolic Care vol. 7 271–277 (2004).

6. Gubbels Bupp, M. R. & Jorgensen, T. N. Androgen-Induced Immunosuppression. Front. Immunol. 9, 794 (2018).

7. Kelly, D. M. & Jones, T. H. Testosterone: a vascular hormone in health and disease. J. Endocrinol. 217, R47–71 (2013).

8. O’Reilly, M. W., House, P. J. & Tomlinson, J. W. Understanding androgen action in adipose tissue. J. Steroid Biochem. Mol. Biol. 143, 277–284 (2014).

9. Davey, R. A. & Grossmann, M. Androgen Receptor Structure, Function and Biology: From Bench to Bedside. Clin. Biochem. Rev. 37, 3–15 (2016).

10. Kaufman, J. M. & Vermeulen, A. The decline of androgen levels in elderly men and its clinical and therapeutic implications. Endocr. Rev. 26, 833–876 (2005).

11. Kaufman, J.-M., Lapauw, B., Mahmoud, A., T’Sjoen, G. & Huhtaniemi, I. T. Aging and the Male Reproductive System. Endocr. Rev. 40, 906–972 (2019).

12. Huhtaniemi, I. & Forti, G. Male late-onset hypogonadism: pathogenesis, diagnosis and treatment. Nature Reviews Urology vol. 8 335–344 (2011).

13. Morgentaler, A. & Rhoden, E. L. Prevalence of prostate cancer among hypogonadal men with prostate-specific antigen levels of 4.0 ng/mL or less. Urology 68, 1263–1267 (2006).

14. Nead, K. T. Androgens and depression: a review and update. Curr. Opin. Endocrinol. Diabetes Obes. 26, 175–179 (2019).

15. Panizzon, M. S. et al. Testosterone modifies the effect of APOE genotype on hippocampal volume in middle-aged men. Neurology 75, 874–880 (2010).

16. Vegeto, E. et al. The Role of Sex and Sex Hormones in Neurodegenerative Diseases. Endocr. Rev. 41, (2020).

17. Vest, R. S. & Pike, C. J. Gender, sex steroid hormones, and Alzheimer’s disease. Hormones and Behavior vol. 63 301–307 (2013).

18. Yeap, B. B. Hormonal changes and their impact on cognition and mental health of ageing men. Maturitas 79, 227–235 (2014).

19. Ruth, K. S. et al. Using human genetics to understand the disease impacts of testosterone in men and women. Nat. Med. 26, 252–258 (2020).

20. Sinnott-Armstrong, N., Naqvi, S., Rivas, M. & Pritchard, J. K. GWAS of three molecular traits highlights core genes and pathways alongside a highly polygenic background. Elife 10, (2021).

21. Prins, B. P. et al. Genome-wide analysis of health-related biomarkers in the UK Household Longitudinal Study reveals novel associations. Sci. Rep. 7, 11008 (2017).

22. Flynn, E. et al. Sex-specific genetic effects across biomarkers. Eur. J. Hum. Genet. 29, 154–163 (2021).

23. Jin, G. et al. Genome-wide association study identifies a new locus JMJD1C at 10q21 that may influence serum androgen levels in men. Hum. Mol. Genet. 21, 5222–5228 (2012).

24. Ohlsson, C. et al. Genetic determinants of serum testosterone concentrations in men. PLoS Genet. 7, e1002313 (2011).

25. Chen, Z. et al. Genome-wide association study of sex hormones, gonadotropins and sex hormone-binding protein in Chinese men. J. Med. Genet. 50, 794–801 (2013).

26. Ellis, L. & Nyborg, H. Racial/ethnic variations in male testosterone levels: A probable contributor to group differences in health. Steroids vol. 57 72–75 (1992).

27. Lopez, D. S. et al. Racial/ethnic differences in serum sex steroid hormone concentrations in US adolescent males. Cancer Causes Control 24, 817–826 (2013).

28. Nyante, S. J. et al. Trends in sex hormone concentrations in US males: 1988-1991 to 1999-2004. Int. J. Androl. 35, 456–466 (2012).

29. Fang, H. et al. Harmonizing Genetic Ancestry and Self-identified Race/Ethnicity in Genome-wide Association Studies. Am. J. Hum. Genet. 105, 763–772 (2019).

30. Bulik-Sullivan, B. K. et al. LD Score regression distinguishes confounding from polygenicity in genome-wide association studies. Nat. Genet. 47, 291–295 (2015).

31. Muntoni, F., Torelli, S. & Ferlini, A. Dystrophin and mutations: one gene, several proteins, multiple phenotypes. Lancet Neurol. 2, 731–740 (2003).

32. Lambert, M. et al. Expression of the transcripts initiated in the 62nd intron of the dystrophin gene. Neuromuscul. Disord. 3, 519–524 (1993).

33. Sironi, M. et al. Transcriptional activation of the non-muscle, full-length dystrophin isoforms in Duchenne muscular dystrophy skeletal muscle. J. Neurol. Sci. 186, 51–57 (2001).

34. Bies, R. D., Friedman, D., Roberts, R., Perryman, M. B. & Caskey, C. T. Expression and localization of dystrophin in human cardiac Purkinje fibers. Circulation 86, 147–153 (1992).

35. Bulik-Sullivan, B. et al. An atlas of genetic correlations across human diseases and traits. Nature Genetics vol. 47 1236–1241 (2015).

36. Zheng, J. et al. LD Hub: a centralized database and web interface to perform LD score regression that maximizes the potential of summary level GWAS data for SNP heritability and genetic correlation analysis. Bioinformatics 33, 272–279 (2017).

37. Giambartolomei, C. et al. Bayesian test for colocalisation between pairs of genetic association studies using summary statistics. PLoS Genet. 10, e1004383 (2014).

38. Grasa, M. D. M. et al. Modulation of SHBG binding to testosterone and estradiol by sex and morbid obesity. Eur. J. Endocrinol. 176, 393–404 (2017).

39. Li, H. et al. Sex Hormone Binding Globulin Modifies Testosterone Action and Metabolism in Prostate Cancer Cells. Int. J. Endocrinol. 2016, 6437585 (2016).

40. Coviello, A. D. et al. A genome-wide association meta-analysis of circulating sex hormone-binding globulin reveals multiple Loci implicated in sex steroid hormone regulation. PLoS Genet. 8, e1002805 (2012).

41. Pereira, A. C. et al. Age and Alzheimer’s disease gene expression profiles reversed by the glutamate modulator riluzole. Mol. Psychiatry 22, 296–305 (2017).

42. Matsuda, S., Matsuda, Y. & D’Adamio, L. BRI3 inhibits amyloid precursor protein processing in a mechanistically distinct manner from its homologue dementia gene BRI2. J. Biol. Chem. 284, 15815–15825 (2009).

43. Carrier, N. et al. The Anxiolytic and Antidepressant-like Effects of Testosterone and Estrogen in Gonadectomized Male Rats. Biol. Psychiatry 78, 259–269 (2015).

44. Nakanishi, Y. et al. Mechanism of Oncogenic Signal Activation by the Novel Fusion Kinase FGFR3-BAIAP2L1. Mol. Cancer Ther. 14, 704–712 (2015).

45. Guo, D.-Q., Zhang, H., Tan, S.-J. & Gu, Y.-C. Nifedipine promotes the proliferation and migration of breast cancer cells. PLoS One 9, e113649 (2014).

46. Errazuriz, I., Dube, S., Basu, A. & Basu, R. Effects of testosterone replacement on glucose and lipid metabolism. Cardiovascular Endocrinology vol. 4 95–99 (2015).

47. Huang, X. et al. VSIG4 mediates transcriptional inhibition of Nlrp3 and Il-1β in macrophages. Science Advances vol. 5 eaau7426 (2019).

48. Helmy, K. Y. et al. CRIg: a macrophage complement receptor required for phagocytosis of circulating pathogens. Cell 124, 915–927 (2006).

49. Vogt, L. et al. VSIG4, a B7 family-related protein, is a negative regulator of T cell activation. J. Clin. Invest. 116, 2817–2826 (2006).

50. Tsukita, S. & Yonemura, S. ERM (ezrin/radixin/moesin) family: from cytoskeleton to signal transduction. Curr. Opin. Cell Biol. 9, 70–75 (1997).

51. Bretscher, A., Edwards, K. & Fehon, R. G. ERM proteins and merlin: integrators at the cell cortex. Nature Reviews Molecular Cell Biology vol. 3 586–599 (2002).

52. Freymuth, P. S. & Fitzsimons, H. L. The ERM protein Moesin is essential for neuronal morphogenesis and long-term memory in Drosophila. Mol. Brain 10, 41 (2017).

53. Li, Y.-Q. et al. Moesin as a prognostic indicator of lung adenocarcinoma improves prognosis by enhancing immune lymphocyte infiltration. World J. Surg. Oncol. 19, 109 (2021).

54. Liao, W., Huang, W., Guo, Y., Xin, M. & Fu, X. Testosterone promotes vascular endothelial cell migration via upregulation of ROCK-2/moesin cascade. Mol. Biol. Rep. 40, 6729–6735 (2013).

55. Malkin, C. J., Pugh, P. J., Jones, R. D., Jones, T. H. & Channer, K. S. Testosterone as a protective factor against atherosclerosis--immunomodulation and influence upon plaque development and stability. J. Endocrinol. 178, 373–380 (2003).

56. Lanser, L. et al. Testosterone Deficiency Is a Risk Factor for Severe COVID-19. Front. Endocrinol. 12, 694083 (2021).

57. Rahim, M. A. et al. Presenilin1 familial Alzheimer disease mutants inactivate EFNB1-and BDNF-dependent neuroprotection against excitotoxicity by affecting neuroprotective complexes of N-methyl-d-aspartate receptor. Brain Communications vol. 2 (2020).

58. Nguyen, A. Q. et al. Astrocytic Ephrin-B1 Controls Excitatory-Inhibitory Balance in Developing Hippocampus. The Journal of Neuroscience vol. 40 6854–6871 (2020).

59. Aqeilan, R. I. et al. Targeted ablation of the WW domain-containing oxidoreductase tumor suppressor leads to impaired steroidogenesis. Endocrinology 150, 1530–1535 (2009).

60. Kunkle, B. W. et al. Genetic meta-analysis of diagnosed Alzheimer’s disease identifies new risk loci and implicates Aβ, tau, immunity and lipid processing. Nat. Genet. 51, 414–430 (2019).

61. Carcaillon, L. et al. Low testosterone and the risk of dementia in elderly men: Impact of age and education. Alzheimers. Dement. 10, S306–14 (2014).

62. Wahjoepramono, E. J. et al. The Effects of Testosterone Supplementation on Cognitive Functioning in Older Men. CNS Neurol. Disord. Drug Targets 15, 337–343 (2016).

63. Jaquish, C. E., Blangero, J., Haffner, S. M., Stern, M. P. & MacCluer, J. W. Quantitative genetics of serum sex hormone—binding globulin levels in participants in the San Antonio Family Heart Study. Metabolism vol. 46 988–991 (1997).

64. Ring, H. Z. et al. Heritability of plasma sex hormones and hormone binding globulin in adult male twins. J. Clin. Endocrinol. Metab. 90, 3653–3658 (2005).

65. Bogaert, V. et al. Heritability of blood concentrations of sex-steroids in relation to body composition in young adult male siblings. Clin. Endocrinol. 69, 129–135 (2008).

66. Shifman, S. Linkage disequilibrium patterns of the human genome across populations. Human Molecular Genetics vol. 12 771–776 (2003).

67. Carrero, J. J. et al. Prevalence and clinical implications of testosterone deficiency in men with end-stage renal disease. Nephrol. Dial. Transplant 26, 184–190 (2011).

68. Snyder, G. & Shoskes, D. A. Hypogonadism and testosterone replacement therapy in end-stage renal disease (ESRD) and transplant patients. Transl. Androl. Urol. 5, 885–889 (2016).

69. Zhao, J. V. & Schooling, C. M. The role of testosterone in chronic kidney disease and kidney function in men and women: a bi-directional Mendelian randomization study in the UK Biobank. BMC Med. 18, 122 (2020).

70. Chen, Y.-X. et al. Role of moesin in renal fibrosis. PLoS One 9, e112936 (2014).

71. Hodonsky, C. J. et al. Genome-wide association study of red blood cell traits in Hispanics/Latinos: The Hispanic Community Health Study/Study of Latinos. PLoS Genet. 13, e1006760 (2017).

72. Carnethon, M. R. et al. Cardiovascular Health in African Americans: A Scientific Statement From the American Heart Association. Circulation 136, e393–e423 (2017).

73. Marshall, M. C., Jr. Diabetes in African Americans. Postgrad. Med. J. 81, 734–740 (2005).

74. Laster, M., Shen, J. I. & Norris, K. C. Kidney Disease Among African Americans: A Population Perspective. Am. J. Kidney Dis. 72, S3–S7 (2018).

75. Nicholas, S. B., Kalantar-Zadeh, K. & Norris, K. C. Racial Disparities in Kidney Disease Outcomes. Seminars in Nephrology vol. 33 409–415 (2013).

76. Norris, K. & Nissenson, A. R. Race, gender, and socioeconomic disparities in CKD in the United States. J. Am. Soc. Nephrol. 19, 1261–1270 (2008).

77. Harris, M. I. et al. Prevalence of Diabetes, Impaired Fasting Glucose, and Impaired Glucose Tolerance in U.S. Adults: The Third National Health and Nutrition Examination Survey, 1988-1994. Diabetes Care vol. 21 518–524 (1998).

78. Bell, R. Diabetes 2001 Vital Statistics. (American Diabetes Association, 2003).

79. Quality, U. S. D. of H. &. H. S. A. F. H. R. A. & US Department of Health & Human Services; Agency for Healthcare Research and Quality. National Healthcare Disparities Report. PsycEXTRA Dataset (2003) doi:10.1037/e308932005-001.

80. Dong, L., Fakeye, O. A., Graham, G. & Gaskin, D. J. Racial/Ethnic Disparities in Quality of Care for Cardiovascular Disease in Ambulatory Settings: A Review. Med. Care Res. Rev. 75, 263–291 (2018).

81. Gibbons, G. H. Physiology, Genetics, and Cardiovascular Disease: Focus on African Americans. The Journal of Clinical Hypertension vol. 6 11–18 (2004).

82. Purcell, S. et al. PLINK: A Tool Set for Whole-Genome Association and Population-Based Linkage Analyses. The American Journal of Human Genetics vol. 81 559–575 (2007).

83. Knight, J. et al. Conditional analysis identifies three novel major histocompatibility complex loci associated with psoriasis. Hum. Mol. Genet. 21, 5185–5192 (2012).

84. Finucane, H. K. et al. Partitioning heritability by functional annotation using genome-wide association summary statistics. Nature Genetics vol. 47 1228–1235 (2015).

85. Gusev, A. et al. Integrative approaches for large-scale transcriptome-wide association studies. Nat. Genet. 48, 245–252 (2016).

86. Benjamini, Y. & Hochberg, Y. Controlling the False Discovery Rate: A Practical and Powerful Approach to Multiple Testing. Journal of the Royal Statistical Society: Series B (Methodological) vol. 57 289–300 (1995).

87. Seabold, S. & Perktold, J. Statsmodels: Econometric and Statistical Modeling with Python. Proceedings of the 9th Python in Science Conference (2010) doi:10.25080/majora-92bf1922-011.

